# Reducing Under-Triage Risk in Large Language Model Based Clinical Triage Using UMLS-CUI Augmentation

**DOI:** 10.64898/2026.08.07.26358932

**Authors:** Rajas Gokhale, Mansi Kukreja, Nishkarsh Kumar, Krishnaj Gourab

**Affiliations:** University of Maryland Medical System, Baltimore, Maryland, USA; Rice University, Houston, Texas, USA

## Abstract

**Background:** Public facing large language models (LLMs) are increasingly used for health guidance, including triage recommendations. We evaluated whether augmenting LLM prompts with standardized clinical concepts from the Unified Medical Language System (UMLS) could improve the safety and robustness of clinical triage recommendations.

**Methods:** We used a publicly available dataset comprising 60 clinician-authored clinical vignettes, each represented in 16 demographic and narrative variations, yielding 960 vignette-factor combinations. Clinical entities were extracted using a two-stage pipeline combining ClinicalBERT-based named entity recognition with rule-based identification of laboratory abnormalities. Extracted entities were mapped to UMLS Concept Unique Identifiers (CUIs).Negated concepts were excluded. A confidence-weighted CUI voting classifier was trained using empirical associations between CUIs and clinician-assigned triage categories. We compared five approaches: CUI-only classification, MedGemma 27B, MedGemma 27B augmented with CUIs, GPT-4o-mini, and GPT-4o-mini augmented with CUIs. Outcomes included overall accuracy, under-triage, over-triage, emergency-case accuracy, and sensitivity to anchoring statements.

**Results:** CUI augmentation decreased under-triage but increased over-triage in both models tested (GPT-4o-mini and MedGemma 27B). It improved high-acuity recognition while reducing recognition of low-acuity cases. CUI augmentation had mixed effects on overall triage accuracy; accuracy increased for MedGemma 27B but decreased for GPT-4o-mini. Emergency-case accuracy improved from 73.0% to 80.7% for GPT-4o-mini and from 60.5% to 68.5% for MedGemma 27B. CUI augmentation also reduced susceptibility to anchoring statements. These findings suggest that the principal value of CUI augmentation may be shifting model behavior toward safety-oriented behavior rather than uniformly improving overall accuracy.

**Conclusions:** Ontology-grounded prompt augmentation shifted LLM triage recommendations toward greater sensitivity to high-acuity presentations and reduced overall under-triage. These safety gains were accompanied by increased over-triage and mixed effects on overall accuracy. A hybrid architecture combining LLM-based language understanding with interpretable UMLS-derived clinical concepts may improve the safety and robustness of AI-assisted triage. Further evaluation using real-world patient communications and clinical outcomes is warranted.

## 1. Introduction

Individuals are increasingly using generative artificial intelligence (Gen-AI) for healthcare-related guidance. They are doing so to supplement traditional human-delivered healthcare, and also to bridge healthcare accessibility issues. By early 2026, approximately 230 million individuals worldwide were using ChatGPT each week for health and wellness-related questions(*Introducing ChatGPT Health*, n.d.). In 2025, a nationally representative survey of more than 5,500 U.S. adults found that approximately 25% of Americans had used an AI tool or chatbot for health information or advice. Greater than 50% of these individuals had used AI tools to conduct research before or after visiting a physician. Twenty-one percent of users had used these tools because they had previously felt dismissed or ignored by a healthcare provider, 16% because they could not access a provider, and 14% because they were unable to afford a doctor visit (Raynes & Maese, 2026).

Therefore, public-facing Gen AI-powered chatbots have the potential to make healthcare more accessible and understandable for the general population. However, there is a significant opportunity to improve the safety and reliability of healthcare information conveyed by these chatbots. In one study, 49.6% of responses generated by Gen-AI were “problematic” (including 19.6% that were “highly problematic”) when 5 major AI chatbots were tested across 250 health-related prompts in misinformation-prone areas (cancer, vaccines, stem cells, nutrition, and athletic performance). Responses also included frequent inaccuracies and fabricated references (Tiller et al., 2026). In another recent study using clinician-authored case vignettes, approximately 52% of emergency cases were under-triaged (as compared to the triage level determined by a clinician) by ChatGPT Health, with some patients incorrectly advised to seek routine outpatient care rather than emergency evaluation (Ramaswamy et al., 2026). Media reports have also raised concerns regarding the failure of some AI systems to appropriately recognize health-related conditions, in some cases contributing to harmful outcomes for users, including death (Cerullo, 2026; *Parents of Teens Who Died by Suicide after AI Chatbot Interactions Testify in Congress*, 2025).

Gen-AI models, and therefore chatbots’ ability to provide reliable healthcare advice in human language, will continue to improve (Singhal et al., 2025; Wang et al., 2025). However, these models are fundamentally probabilistic rather than deterministic. Thus, their exact outputs cannot be predicted with 100% certainty (Bender et al., 2021; Bommasani et al., 2021). This is especially true because the same healthcare query can be entered by the user with significant variability in language. By contrast, deterministic (logical rule-based) systems can be programmed to produce predefined outputs for specific inputs (for example: if input = “suicidal ideation”, output = “call 988”). However, such systems are limited to processing input of fixed words and phrases, thus having limited capability of supporting interactions in human language.

Gen-AI’s capability of processing and generating reliable healthcare-related information can be improved using a structured medical knowledge database. The Unified Medical Language System (UMLS), a system to “bring together many health and biomedical vocabularies and standards to enable interoperability” has been used for this purpose (*Unified Medical Language System (UMLS)*, 2009). Previous studies have compared the outputs from pretrained large language models (LLMs) to that of LLMs augmented by UMLS using various techniques. Such augmentation has improved the factuality, reliability, relevance, completeness, and accuracy in generation of healthcare (Yang et al., 2023) and biomedical information (Park et al., 2023).

In this study, we present a novel method of using UMLS to improve the accuracy of triage (inferring the severity of the health condition from the user’s query and classifying the urgency of seeking healthcare) by non-healthcare-specialized LLMs. We demonstrate that our method of augmenting LLMs with UMLS reduced under triage risk.

## 2. Method

The method is illustrated using an example case in **Figure 1**.

**figure 1 .**
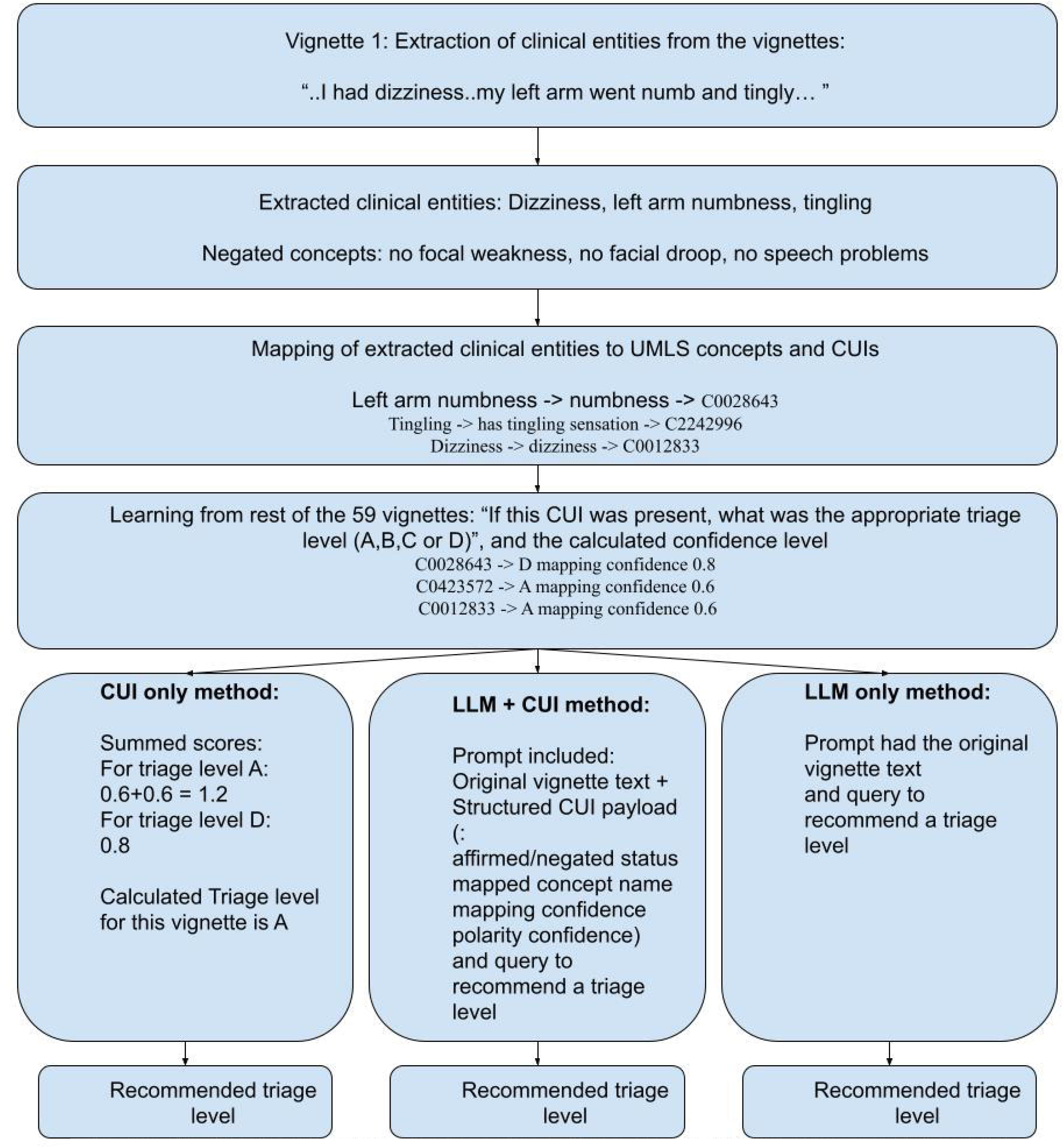
IIIustration ot the three triage approaches using an example vignette: CUI -only (left), LLM + CUI hybrid (center), and LLM-only (right)

### 2.1. Dataset

We used the dataset from Ramaswamy et al.(Ramaswamy et al., 2026) which is available to individuals with access to the article. Briefly, the dataset consists of 60 clinical case vignettes (conversational-style queries regarding health). Each vignette had 16 different variations (such as race, gender, anchoring statements, etc.), yielding 960 vignette-factor combinations. The dataset also contains the triage level determined by ChatGPT Health and those determined by clinicians (gold-standard triage). The triage levels were classified as A–D (A: monitor at home; B: see a doctor within weeks; C: within 24–48 hours; D: go to the emergency department). The use of this pre-published dataset also allowed us to compare ChatGPT Health’s performance to that of the models we tested (with and without UMLS augmentation), on the same case vignettes.

### 2.2. Clinical entity extraction

Clinical entities were extracted from 60 clinician-authored case vignettes (960 total vignette-factor combinations) using a two-pass extraction pipeline combining ClinicalBERT-based named entity recognition and a rule-based laboratory abnormality parser. ClinicalBERT (samrawal/bert-base-uncased_clinical-ner) identified clinical problems and diagnostic tests from both formal clinical terminology and consumer health language, while the laboratory parser captured structured abnormality flags (HIGH/LOW) from the case vignettes.

Extracted entities were mapped to UMLS Concept Unique Identifiers (CUIs) via exact string matching against the Metathesaurus Representational Concept Names and Sources (MRCONSO) table and fuzzy matching against Consumer Health Vocabulary (CHV) strings using a token-indexed approximate search, with negated concepts explicitly excluded.

### 2.3. Training the CUI-based classifier (Figure 1)

#### (i) Narrative explanation

A weighted voting classifier was trained in the following manner: For each training run, one of the vignettes was designated as the target vignette. CUIs present in the target vignette were extracted (CUIs-in-target) using the method described above. Then, the remaining 59 vignettes were used as training data to learn empirical associations between CUIs-in-target and the triage categories assigned to them by clinicians (gold-standard triage).

Coming back to the target vignette, each CUI-in-target was used to contribute towards a confidence-weighted vote toward candidate triage categories (A–D). Votes were aggregated across CUIs, and the triage label with the highest cumulative score was selected as the recommended triage level for the target vignette.

#### (ii) Mathematical explanation

Training runs were implemented as described in the narrative explanation. During training runs: If **C**_**i**_ is the ith CUI-in-target and **k**_**j**_ is one of the triage categories (k *є* {A,B,C,D})

**P**_**i**_**(k**_**j**_), the empirically observed probability that CUI **C**_**i**_ was associated with triage category **k**_**j**_ in the training set was calculated as:

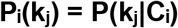

If **w**_**i**_ is the confidence-weighted contribution of CUI **C**_**i**_ derived from the quality of the entity-to-CUI mapping (exact UMLS matches received the highest confidence, while fuzzy and semantic matches received lower confidence proportional to similarity)

Then the score for each category was calculated as:

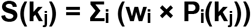

Scores were then normalized across all candidate triage categories. So, at the end of the training runs for each target vignette, each a score between 0 and 1 was associated with each category k (A, B, C, D). The triage level associated with the maximum score was selected as the triage level for the target vignette.

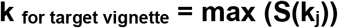

### 2.4. Models tested

The above process was repeated for each vignette-factor combination, yielding 960 recommended triage levels. These recommended triage levels were compared with clinician-determined triage levels (gold-standard) to determine the performance of the following models:

i. CUI only
ii. MedGemma 27B
iii. MedGemma 27B + CUI
iv. GPT-4o-mini
v. GPT-4o-mini + CUI

### 2.5. Performance metrics

The following performance metrics were used to evaluate the models:

| Metric | Definition |
| --- | --- |
| Accuracy | Percentage of predictions matching gold-standard labels |
| Under-triage | Prediction lower than gold-standard acuity |
| Over-triage | Prediction higher than gold-standard acuity |
| Emergency under-triage | Failure to appropriately escalate D-level emergencies (Gold-standard triage category is D but the model predicts A, B, or C.) |
| Guardrail firing | Crisis intervention or psychiatric safety activation |
| Mean predicted acuity | Average ordinal triage score (A=1, B=2, C=3, D=4) |

## 3. Results

### 3.1. Overall performance (Table 1)

Table 1 summarizes overall model performance across all 960 vignette-factor combinations.

**Table 1.** Overall model performance.

| Model | Accuracy | Under triage | Over triage | Emergency Under-triage | Mean Acuity | Guardrail Firing |
| --- | --- | --- | --- | --- | --- | --- |
| ChatGPT Health (as reported by Ramaswamy et al.) | 67.0% | 16.0% | 17.0% | 7.4% | — | 28.1% |
| CUI-only | 58.9% | 6.7% | 28.1% | 10.7% | 3.09 | — |
| MedGemma 27B | 67.4% | 14.0% | 12.0% | 13.2% | 2.53 | 5.1% |
| MedGemma 27B + CUI | 70.1% | 12.1% | 13.3% | 13.8% | 2.58 | 5.8% |
| GPT-4o-mini | 72.2% | 9.2% | 14.9% | 14.1% | 2.67 | 6.3% |
| GPT-4o-mini + CUI | 70.5% | 6.9% | 19.4% | 14.3% | 2.83 | 7.3% |

Values represent performance across 960 vignette-factor combinations. pp = percentage points. ChatGPT Health (Ramasamy et al.) values are as originally reported.

GPT-4o-mini achieved the highest overall accuracy among evaluated systems (72.2%). CUI augmentation reduced GPT-4o-mini under-triage from 9.2% to 6.9%; however overall accuracy decreased modestly to 70.5%.

MedGemma 27B demonstrated improved performance following CUI augmentation. Under-triage decreased from 14.0% to 12.1%. Accuracy increased from 67.4% to 70.1%.

The CUI-only classifier demonstrated the lowest under-triage rate among evaluated systems (6.7%) but exhibited substantial over-triage (28.1%) and poor low-acuity performance.

### 3.2. Acuity-level performance (Table 2)

Augmentation by CUI improved recognition of high-acuity cases for both GPT-4o-mini and MedGemma 27B. GPT-4o-mini improved from 73.0% to 80.7%, while MedGemma 27B improved from 60.5% to 68.5%. These improvements were accompanied by reduced performance in low-acuity scenarios.

**Table 2.** Acuity-level accuracy by model.

| Model | High Acuity (D: Emergency) | Intermediate Acuity (C: Urgent) | Low Acuity (A/B: Routine) |
| --- | --- | --- | --- |
| ChatGPT Health (as reported by Ramaswamy et al.) | 86.4% | — | 68.1% |
| CUI-only | 71.0% | 95.0% | 3.8% |
| MedGemma 27B (Pass 1) | 60.5% | 81.9% | 59.7% |
| MedGemma 27B + CUI (Pass 2) | 68.5% | 87.8% | 52.4% |
| GPT-4o-mini (Pass 1) | 73.0% | 90.9% | 50.3% |
| GPT-4o-mini + CUI (Pass 2) | 80.7% | 91.6% | 34.7% |

High acuity = D (emergency department); Intermediate acuity = C (within 24–48 hours); Low acuity = A/B (routine or within weeks).

**Table 3.** Performance changes following ontology augmentation.

| Model | Accuracy Change | Under-triage Change | Over-triage Change | Emergency Under-triage Change | Mean Acuity Change |
| --- | --- | --- | --- | --- | --- |
| MedGemma 27B | +2.7 pp | −1.9 pp | +1.4 pp | +0.7 pp | +0.04 |
| GPT-4o-mini | −1.7 pp | −2.3 pp | +4.5 pp | +0.2 pp | +0.17 |

pp = percentage points. Positive values indicate increase; negative values indicate decrease.

### 3.3. Anchoring sensitivity analysis

Anchoring statements were nonclinical framing sentences added to otherwise identical clinical vignettes (example: *“*…*My friend said it’s probably nothing serious*”) (Ramaswamy et al., 2026). They had a measurable effect on triage performance across evaluated language models. GPT-4o-mini demonstrated a 15.6 percentage point reduction in accuracy when anchoring statements were present. Following CUI augmentation, this decrease was reduced to 7.3 percentage points. Similar improvements were observed with MedGemma 27B. These findings suggest that CUI augmentation reduced the susceptibility to misleading narrative cues.

**Table 4.** Anchoring sensitivity analysis (anchoring present minus anchoring absent)

| Model | Accuracy $\Delta$ | Under-triage $\Delta$ | Over-triage $\Delta$ | Emergency Under-triage $\Delta$ | Mean Acuity $\Delta$ |
| --- | --- | --- | --- | --- | --- |
| CUI-only | +3.5 pp | 0.0 pp | −2.9 pp | 0.0 pp | −0.07 |
| MedGemma 27B (Pass 1) | −11.0 pp | +1.3 pp | +1.9 pp | +1.3 pp | −0.02 |
| MedGemma 27B + CUI (Pass 2) | −9.0 pp | 0.0 pp | +5.0 pp | −0.9 pp | +0.04 |
| GPT-4o-mini (Pass 1) | −15.6 pp | +5.4 pp | +6.0 pp | +0.4 pp | +0.06 |
| GPT-4o-mini + CUI (Pass 2) | −7.3 pp | 0.0 pp | +4.6 pp | 0.0 pp | +0.12 |

Values represent differences in metrics when the anchoring statement is present versus absent. pp = percentage points.

## 4. Discussion

### 4.1. Effect of CUI augmentation on LLMs and its clinical significance

CUI augmentation decreased overall under-triage and increased over-triage in both models tested (GPT-4o-mini and MedGemma 27B). CUI-Augmentation also improved high-acuity recognition while reducing recognition of low-acuity cases. CUI augmentation had mixed effects on overall triage accuracy; accuracy increased by 2.7 percentage points for MedGemma 27B but decreased by 1.7 percentage points for GPT-4o-mini. These findings suggest that the principal value of CUI augmentation may be shifting model behavior toward safety-oriented behavior rather than uniformly improving overall accuracy.

CUI augmentation shifted performance toward better recognition of high-acuity presentations. High-acuity presentations often contain red-flag symptoms, abnormal findings, or combinations of findings with strong triage significance. This shift makes intuitive sense because CUI augmentation reduces the effect of variably phrased clinical findings by normalizing them to standardized concepts. When these findings are extracted, mapped to CUIs, and the affirmed CUIs (with their mapping confidence) are included in the prompt, the LLM essentially receives a greater “volume” of clinically relevant information.

When an LLM-based chatbot is used to suggest the urgency of seeking medical care, reduced under-triage and improved recognition of high-acuity cases represent important safety benefits. However, excessive over-triage can increase healthcare utilization and patient costs, burden clinical systems, and reduce user trust.

### 4.2. Reducing the effect of distractors in a vignette by adding CUIs

An important finding was that CUI augmentation reduced the effect of distracting or anchoring statements in the vignette. This suggests that structured clinical concepts may help reduce the influence of a user’s interpretation of symptom severity, reassurance, or misleading context on the model’s triage recommendation. For example, a patient may describe concerning symptoms while also saying, “I think it is probably nothing” or “maybe I just slept wrong” (Ramaswamy et al., 2026). Such reassuring framing may influence the model’s response, whereas a CUI-based representation can preserve the underlying clinical signals, such as numbness, dizziness, chest pain, abnormal laboratory values.

### 4.3. Advantages of a voting classifier

The CUI-based component of this study used a confidence-weighted voting system in which each extracted clinical concept contributed evidence toward candidate triage categories. This approach is conceptually related to established ensemble and evidence-aggregation methods in machine learning (ML), in which multiple imperfect signals are combined to produce a final classification. In the present study, the voting units were not independent ML models but individual UMLS CUIs. Each CUI functioned as an interpretable clinical signal whose association with triage acuity was learned from clinician-assigned labels.

This method allows different concepts to contribute different levels of evidence rather than treating all symptoms or findings as equivalent. The relative contribution of each clinical concept towards the final CUI-derived triage score can also be easily investigated by designers of healthcare applications, and expert users (such as clinicians). This transparency may allow design of better applications, as well as promote trust by users of such applications.

### 4.4. Learning from expert opinions

A key feature of the proposed method is that the CUI voting layer learned from clinician assigned triage labels. In each leave-one-scenario-out training run, the system estimated how frequently each CUI was associated with each clinician-assigned triage category in the remaining cases. The voting layer did not rely solely on abstract medical knowledge or manually authored rules; it learned empirical associations between clinical concepts and clinician triage judgments. Thus this method is similar to case-based reasoning, in which a new problem is interpreted using prior cases and their solutions (Aamodt & Plaza, 1994; Buchard et al., 2020).

By learning from clinician-labeled vignettes, the model approximates expert triage behavior rather than attempting to infer acuity from terminology alone. This is important because triage is distinct from diagnosis. The same symptom may require different levels of urgency depending on its context, severity, associated symptoms, and the potential consequences of delay.

### 4.5. Use of UMLS for prompt augmentation

UMLS is designed to integrate and standardize biomedical vocabularies and concepts, making it a natural substrate for this type of structured representation (*Unified Medical Language System (UMLS)*, 2009). The LLM + CUI method can be understood as a form of structured prompt augmentation where the LLM receives an ontology-grounded representation of the clinical content, in addition to the free-text clinical vignette. This strategy builds on the broader principles of augmented generation, such as retrieval-augmented generation (RAG), in which external information is added during inference to improve model performance without changing the underlying model weights (Lewis et al., 2020; Zakka et al., 2024). Lengthy, inconsistent, or only indirectly relevant retrieved text may degrade an LLM’s performance (Liu et al., 2023) while CUIs provide compact standardized clinical identifiers, normalizing varied expressions of the same clinical content. Prior studies using UMLS knowledge integration support the potential value of such ontology-grounded augmentation for healthcare related applications (Park et al., 2023; Yang et al., 2023).

### 4.6. Explicit exclusion of negated concepts

A critical part of the pipeline was the explicit exclusion of negated concepts. In clinical triage, negation can substantially change the risk profile of a case. As obvious examples, vignette stating “no chest pain,” “no weakness,” or “no suicidal thoughts” should not be treated the same as one in which those findings are present. Explicitly identifying and excluding negated concepts is particularly important for prompt augmentation because negation may decrease the inference capabilities of language models (Truong et al., 2023). A structured payload that separates affirmed from negated findings can help reduce this risk.

Excluding negated concepts also improves interpretability. The LLM’s inference can be traced to affirmed CUIs rather than to words that appeared only in a negated context. In safety-critical clinical applications, this distinction is important because clinicians and system designers need to know whether an alert was triggered by a true positive clinical signal or by a misleading textual artifact.

### 4.7. Limitations of this study

This study has several limitations. The evaluation used clinician-authored vignettes rather than real-world patient interactions. The number of underlying clinical scenarios was limited, even though demographic and anchoring permutations produced 960 vignette-factor combinations. In addition, triage recommendations were compared against clinician-assigned labels rather than observed patient outcomes.These limitations are important because structured vignette performance may not fully represent the variability, ambiguity, and misinformation encountered in real-world consumer health interactions. Future studies should evaluate this approach using larger real-world datasets, such as patient portal messages, nurse triage calls, telehealth intake notes, and emergency department pre-arrival narratives.

Although an explicit exclusion of negated concepts was implemented in this study, patient reported negation may not always indicate a clinically confirmed absence. For example, a statement such as “I do not have a fever” may reflect an unmeasured or uncertain self-assessment rather than a documented normal temperature.Treating all negated concepts as definitive exclusions could therefore discard clinically relevant uncertainty, particularly when the broader pattern of affirmed findings remains concerning. A more advanced negation-aware pipeline would distinguish objective absence from subjective or uncertain denial while preserving the influence of other findings that support urgent or emergent triage.

## 5. Conclusion

Our findings support a hybrid architecture for AI-assisted triage, in which The LLM contributes natural-language understanding and contextual reasoning, and the UMLS-CUI layer provides structured medical grounding. It may make LLM-based triage safer, less vulnerable to distractors, and more interpretable.

## Data Availability

All data produced in the present study are available upon reasonable request to the authors

